# Investigating sensitivity of nasal or throat (ISNOT): A combination of both swabs increases sensitivity of SARS-CoV-2 rapid antigen tests

**DOI:** 10.1101/2022.01.18.22269426

**Authors:** Barbara L Goodall, Jason J LeBlanc, Todd F Hatchette, Lisa Barrett, Glenn Patriquin

**Affiliations:** Department of Medicine, Dalhousie University, Halifax, Nova Scotia, Canada; Department of Pathology, Dalhousie University, Halifax, Nova Scotia, Canada; Division of Microbiology, Department of Pathology and Laboratory Medicine, Nova Scotia Health, Halifax, Nova Scotia, Canada; Department of Microbiology and Immunology, Dalhousie University, Halifax, Nova Scotia, Canada

**Keywords:** COVID-19, SARS-CoV-2, rapid, antigen, nasal, throat

## Abstract

The COVID-19 pandemic has been hallmarked by several waves of variants of concern (VoCs), each with novel challenges. Currently, the highly transmissible Omicron VOC is predominant worldwide, and sore throat is common among other cold-like symptoms. Anecdotes on social media suggested sampling one’s throat can increase sensitivity for Omicron detection by antigen-based rapid testing devices (Ag-RDTs). This work determines whether the sensitivity of Ag-RDTs designed for nasal sampling is altered with use of self-administered throat swabs in self-perceived asymptomatic individuals. This investigation compared results of a common Ag-RDT (i.e. Abbott Panbio COVID-19 Ag Rapid Test Device) using three sampling sites: nasal swab; throat swab and; combined nasal/throat. All Ag-RDT results were confirmed with molecular testing. Compared to RT-PCR, samples from nasal or throat swabs each detected 64.5% of SARS-CoV-2 cases; however, combining the contributions of each swab increased sensitivity to 88.7%. This trend was also evident with the Rapid Response Ag-RDT (BTNX), which uses a more flexible swabs than Panbio. When nasal swab collection was compared to paired sampling of the nasal/throat using a single swab with the Panbio Ag-RDT, the sensitivity of each was 68.4% and 81.6%, respectively. No false-positive results were observed with nasal, throat, or combined nasal/throat sampling. Self-administered throat and nasal/throat swabs both had >90% acceptability. These findings support the use of self-collected combined nasal/throat sampling for Ag-RDT based SARS-CoV-2 detection in self perceived asymptomatic individuals.

## Introduction

The COVID-19 pandemic is an ongoing threat to global public health. Since the first cases were reported in December 2019, several SARS-CoV-2 variants concern (VoCs) have emerged, causing multiple waves of infection globally^1^. In the last 4 weeks, the predominant VoC in many jurisdictions is SARS-CoV-2 lineage B.1.1.529, designated Omicron by the World Health Organization (WHO) classification working group^1^, which is highly mutated, with over 50 mutations compared to ancestral reference genomes^2,3^. Most mutations are present in the gene encoding spike, the protein that is targeted by all current vaccines, and plays a role in host receptors interactions to facilitate viral entry and subsequent replication. Among other Omicron mutations, four mutations are in the gene encoding nucleocapsid, the target of most antigen-based rapid diagnostic tests (Ag-RDTs). While some recent data have alleviated initial concerns over the possibility of decreased sensitivity of Ag-RDTs with Omicron, there are other noted differences from previous VOCs^4^. Omicron has increased transmission, reduced vaccine effectiveness^5,6^, and recent studies have suggested there could be differences in tissue tropism^3,7,8^.

Clinically prominent cold-like symptoms, especially marked sore throat^9^, combined with reports of perceived decreased sensitivity of self-performed Ag-RDTs with bilateral nasal swab collection, have led to anecdotal recommendations to self-swab one’s throat (with or without nasal swabbing), contradicting manufacturer directions. These views have been widely propagated on social media, and been discussed and debated in recent print and televised media^10–12^. From a virus perspective, differences in viral tropism and kinetics of Omicron may be different compared to previous variants^7,8^, and adds some credence to the idea that virus may be in different parts of the respiratory system at different times.

Many factors can affect Ag-RDT sensitivity, including the timing and quality of specimen collection, the specimen type, and the testing method itself^13,14^. Previous work has also shown that deviating from the manufacturer’s instructions for rapid antigen tests can introduce errors that may yield inaccurate results^15^. Misunderstanding erroneous test results can fuel misinformation and disinformation, ultimately undermining confidence in public health efforts and trust in healthcare professionals.

At a low barrier volunteer-led community testing centre^16^, whereby samples were self-collected with coaching, this work sought: to investigate whether Omicron could be detected with Ag-RDTs in asymptomatic individuals; to evaluate the potential benefits of throat sampling; and assess the applicability of throat sampling to other Ag-RDTs using different swabs.

## Methods

### Sample collection and SARS-CoV-2 Ag-RDTs

Participants were community members attending an urban rapid testing location available specifically for those who self-identified as being asymptomatic^16^, over a seven-day period in January 2022. After providing verbal consent, participants were verbally instructed on, and observed performing, a self-collected bilateral nasal swab, and then guided on self-collection of a posterior oropharyngeal swab, with the aid of an anatomical diagram of the mouth. In a second series of evaluations, participants were instructed on self-swabbing for a bilateral nasal sample, followed by a combined throat/bilateral nasal sample. All swabs were collected by the participants with minimal coaching by trained volunteers using the Panbio COVID-19 Ag Rapid Test Device (Abbott Rapid Diagnostics, Jena, Germany) according to the manufacturer’s instructions. Swabs included with other Ag-RDTs differ in wand flexibility, and may have different characteristics in throat sampling. Therefore, for a sub-set of participants with a positive Panbio Ag-RDT (from either anatomical site), individual self-collected swabs from the anterior nares bilaterally and throat were repeated and processed using the Rapid Response COVID-19 Antigen Rapid Test Device (BTNX Inc., Markham, ON). All Ag-RDTs were interpreted by volunteer testing staff according to manufacturer instructions, with a visualized test (T) band considered positive (in conjunction with a visualized control (C) band), regardless of intensity (Figure). For supplemental results, positive reaction bands were graded according to intensity relative to the control band, whereby an intensity equal to that of the control band was deemed “++”, with less-intense bands as “+” and more-intense bands graded as “+++”. Bands that were barely visible, were graded as “+/-”.

**Figure.**
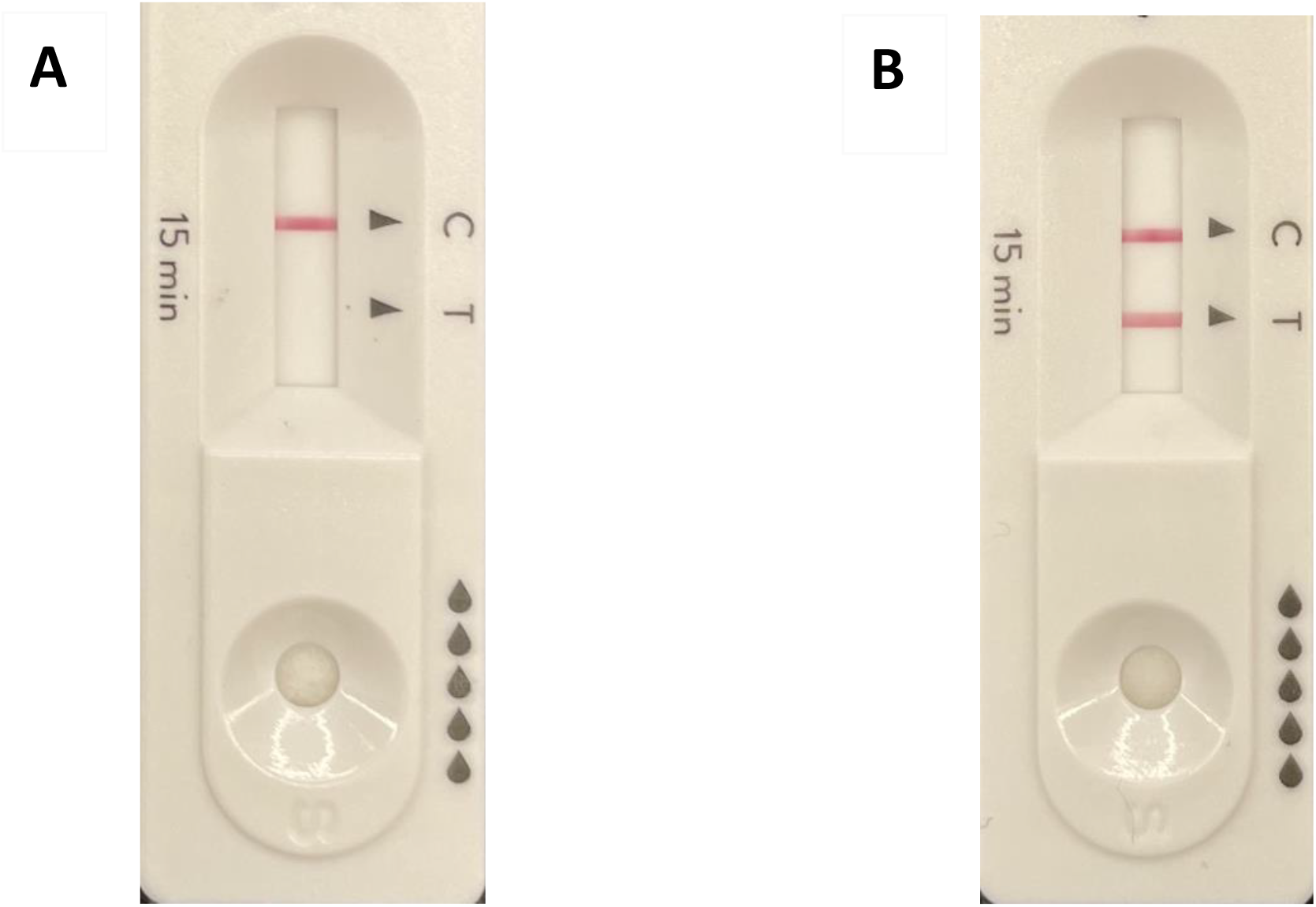
Representative example of a single participant’s results using the Abbott Panbio antigen-based rapid testing device, for (A) nasal sampling alone, and (B) combined nasal/throat sampling. Bands represent the control line (C) and test line (T), indicating a negative result for A and the detection of SARS-CoV-2 for B.

### SARS-CoV-2 RT-PCR

Real-time RT-PCR was performed on residual test buffer (RTB) from the Ag-RDT kits to confirm positive antigen detections and to investigate the possibility of false negative results^16^. Briefly, the remaining fluid in the Ag-RDT tubes along with the used swabs for all participants (Ag-RDT positive or negative) were transported to a central laboratory, and 200 µl of viral transport medial (VTM) (Rodoxica, Little Rock AR) was added to each tube. Following gentle vortexing for 5 seconds, the VTM/fluid was transferred to a microcentrifuge tube, and 150 µl was subjected to a total nucleic acid (TNA) extraction on a MaNAPure LC 2.0 instrument (Roche Diagnostics ltd., Roltkreuz, Switzerland), according to manufacturer instructions. TNAs were eluted in a volume of 50 µl and 5 µl was used as template in a real-time RT-PCR using the TaqPath COVID-19 Combo Kit (Life Technologies Corp., Frederick, MD) according to the manufacturer’s directions.

### Data analysis

Results were defined at the level of the participant, using RT-PCR as the reference standard. Any antigen-positive result from any individual’s swabs confirmed by RT-PCR was considered positive. A positive Ag-RDT result that failed to confirm with RT-PCR testing was interpreted as a false positive. False negative Ag-RDT results were defined by detection of SARS-CoV-2 by RT-PCR in absence of an Ag-RDT positive result. Concordant negative results by both Ag-RDT and RT-PCR were considered negative. Descriptive statistics were used to report participation and testing outcomes, using MedCalc (http://www.medcalc.org) where appropriate. Sensitivity and specificity were reported with 95% confidence intervals (CI) for all investigations, with the exception of specificity for BTNX, which was not calculated due to the lack of appropriate denominator.

## Results

### Comparison of nasal and throat samples using Panbio Ag-RDT

A total of 1568 people attended the testing site during the seven-day investigation period, of whom 1472 (93.9%) consented to participate. Of those who participated in the phase of separate bilateral nasal and throat swabbing, 40 positive Panbio Ag-RDT results each were obtained with both the nasal and throat swab, out of 62 RT-PCR-positive individuals. As such, the sensitivity for nasal or throat swabs was identical at 64.5% [95% confidence interval (CI): 52.1 to 75.3%] (Table). When detections attributed to either swab were combined in the same analysis, 55 of 62 individuals were identified as positive by Panbio Ag-RDT, for a combined sensitivity of 88.7% [95% CI: 78.2 to 94.7%] (Table). No false positives were observed, and as such, the specificity of each swab was 100% [95% CI: 99.5 to 100.0%] (Table). Of note, all RT-PCR-positive samples featured an S-gene drop-out (Supplemental Tables), which is consistent with the circulating Omicron VOC.

**Table.** Sensitivity and specificity of sampling different anatomical sites for SARS-CoV-2 detection by antigen-based rapid detection tests.

| Test | Swab type | Sensitivity | 95% CI | Specificity | 95% CI |
| --- | --- | --- | --- | --- | --- |
| **Abbott Panbio** | Nasal alone | 64.5% | 52.1 to 75.3% | 100.0% | 99.5 to 100.0% |
|  | Throat alone | 64.5% | 52.1 to 75.3% | 100.0% | 99.5 to 100.0% |
|  | Combined results | 88.7% | 78.2 to 94.7% | 100.0% | 99.5 to 100.0% |
| **BTNX Rapid Response** | Nasal alone | 78.1% | 62.4 to 89.4% | NA | NA |
|  | Throat alone | 63.4% | 46.9 to 77.9% | NA | NA |
|  | Combined results | 95.1% | 83.5 to 99.4% | NA | NA |
| **Abbott Panbio** | Nasal alone | 68.4% | 51.4 to 82.5% | 100.0% | 99.2 to 100.0% |
|  | Combined nasal + throat swab | 81.6% | 65.7 to 92.3% | 100.0% | 99.2 to 100.0% |

CI, confidence interval; NA, not applicable

### Nasal and throat swabs comparison using the BTNX Ag-RDT

To verify the generalizability of results of the Panbio Ag-RDT results, a subset of participants with a positive Panbio Ag-RDT was recruited for additional testing with repeat nasal and throat swabs, which were then subjected to the BTNX Rapid Response Ag-RDT. This particular assay was chosen given that the swab packaged with testing device is a thin, flexible, flocked swab suitable for nasopharyngeal swabbing, in contrast to the thick, rigid swabs packaged in most Ag-RDTs (like the Panbio assay). The nasal swab was positive in 32/41 participants (78.1% [95% CI: 62.4 to 89.4%]) and the throat swab was positive in 26/41 participants (63.4% [95% CI: 46.9 to 77.9%]), but with results combined, 39/41 (95.1% [95% CI: 83.5 to 99.4%]) of cases were identified. No false-positive BTNX swabs were observed.

### Evaluation of a combined nasal/throat swab using Panbio

After confirming the sensitivity of throat and nasal swabs alone, the performance characteristics of a combined nasal/throat swab was evaluated to facilitate the practical applications of Ag-RDT sampling both anatomical sites. For this phase of the quality project, participants were asked to provide a combined nasal/throat swab after having completed the standard bilateral nasal swab, and each swab was subjected to the Panbio Ag-RDT. A total of 520 individuals participated, of whom, 38 were positive by either swabbing method. Bilateral nasal swabs resulted in 26 positive Ag-RDTs, with a sensitivity of 68.4% [95% CI: 51.4 to 82.5%]. In comparison, combined nasal/throat swabbing identified 31 positive participants (Figure), for a sensitivity of 81.6% [95% CI: 65.7 to 92.3%] (Table). As no false positives were detected in the evaluation period, a specificity of 100% [95% CI: 99.2 to 100.0%] was obtained for both swabs.

## Discussion

A global crisis such as a pandemic and the rapid dissemination of information through social media can fuel perpetuation of anecdotes, misinformation, and disinformation that can hamper public health efforts and trust in healthcare professionals. This quality project assessed assertions from media (social, print, televised), urging the public to swab their throat in addition to (or instead of) their nares for testing with Ag-RDTs. In this project, a combined collection of both nasal and throat samples increased Ag-RDT sensitivity compared to the manufacturer recommended sampling of bilateral nares alone. To our knowledge, this is the first report of the self-collected Ag-RDT performance of Ag-RDTs for Omicron detection in an asymptomatic population.

Given the propensity of cold-like symptoms and complaints of sore throat during this period of high prevalence of Omicron, there was likely some validity to the media accounts of false-negative nares swabs, with subsequent follow-up positive throat swabs. Many of the Ag-RDTs available to the public are based on nasal swabs, and are not approved for sampling saliva or the throat and previous work shows the potential for invalid results when using an assay outside of the manufacturer’s instructions^15^. Without careful evaluation of this widely publicized phenomenon, variables such as test timing (and time between nasal and throat tests), testing kit manufacturer, swab types, and buffers, cannot be controlled. Importantly, confirmation of test results with more sensitive and specific assays (such as RT-PCR) cannot be easily performed to corroborate individual anecdotes. Investigations in this work allowed for careful evaluation of SARS-CoV-2 detection using Ag-RDTs, using nasal and throat swabs.

Prior to undertaking this evaluation, one of the primary concerns for the wide use of throat-based collection for Ag-RDT testing was the potential for false positives. While food and beverages could potentially impact the performance of Ag-RDTs or of molecular testing^17^, all of the Ag-RDT-positive throat samples in this project were confirmed as positive by RT-PCR. Based on our experience with community-based testing over the last year, the estimated false positive rate was between 2 and 4 per 1000 individuals, which is consistent with manufacturer claims. Given the widespread activity of Omicron in the community, it was not surprising that few false positives were observed.

Apart from ensuring Ag-RDT specificity with use of a throat-based collection, understanding its potential impacts on sensitivity is also crucial. The sensitivity of the Ag-RDT for Omicron detection was 82% (versus RT-PCR) when using the combined nasal and throat swabs, which is on the higher end of estimates previously reported for both Omicron and non-Omicron variants. The most recent Cochrane review of the performance of Ag-RDTs prior to the Omicron wave suggested the sensitivity of the Panbio assay was between 50% and 92%^18^. Although the US Food and Drug Administration (FDA) recently issued a statement that antigen test may have reduced sensitivity for the detection of Omicron^19^, real-world data was lacking. Consistent with others^4,20^, the National Microbiology Laboratory (NML) in Canada recently assessed the performance of several Ag-RDTs for Omicron detection, and results for Panbio and BTNX were comparable to previous evaluations using other SARS-CoV-2 lineages (NML unpublished communication).

The added detection from sampling the throat could be due to several factors. Sampling error or less vigorous sampling of one site versus the other would influence the quality of the specimen. With the use of RTB following Ag-RDT testing, molecular testing could be performed to understand SARS-CoV-2 cases that would be potentially missed by Ag-RDTs using nasal and throat swab-based collections. As expected for any Ag-RDT, there were false negative Ag-RDT results detected by RT-PCR, but these were equally distributed between the nasal and throat-based collections. It is also possible that the discrepancy between nasal and throat results is attributed to preferential replication of the Omicron variant in these anatomical sites at different time points in infection. Prior to the first descriptions of Omicron, reports of RT-PCR from comparisons of nasopharyngeal and oropharyngeal/nares^21^, and nasal and oropharyngeal^22^ swabs showed little difference. On the other hand, in symptomatic non-hospitalized patients in South Africa, saliva samples detected Omicron more often than mid-turbinate swabs (100% vs. 86% respectively)^8^ which was opposite from the performance reported for previously circulating variants. Similarly, in a pre-print non-peer reviewed publication of a small subset of individuals in a high-risk occupational case cohort, the viral load in saliva specimens peaked 1 to 2 days prior to those observed with nasal swab collections^23^. These early data suggest there may be a different tissue tropism for the Omicron variant compared to prior circulating lineages of SARS-CoV-2^7^.

Most individuals being tested agreed to participate, with only approximately 6% refusing, indicating that a self-administered throat swab is an acceptable method for COVID-19 testing. Tolerability of throat swabs and of combined nasal/throat swabs appeared to be high, without voiced concerns for swab preference. Participants often performed the self-administered throat swab correctly with minimal coaching. Many participants experienced a gag reflex during the self-administered throat swab, and one individual vomited. This in-person volunteer observation and coaching could easily be translated to a brief instructional video for at-home/occupational use.

This investigation is not without limitations. Our adult population is from a single centre during a seven-day period, at a community testing centre. There was no collection of clinical information related to patient demographics, timing from exposure, immunization history, or recent consumption of food or drink. One of the criteria for attending the testing site was self-described asymptomatic status. However, mild symptoms may not have been recognized as Omicron infection. Practically speaking, swabbing both nasal and throat using a single swab can be done without regard for timing of symptoms or exposure, and should increase rates of detection using Ag-RDTs. This is particularly important as self-directed testing and management become a tool of endemic living, these data add confidence to the ability of individuals to adequately perform reliable sample collection in a real-world setting.

Although this project focused on the Abbott Panbio Ag-RDT, the concordant results obtained using the BTNX Rapid Response suggest that this enhanced detection rate using a combined swab is applicable to other lateral flow tests, although further studies are warranted. While molecular confirmation with RT-PCR was available, RTB is not a typical testing matrix. Despite our previous validations of RTB for this application^16^, this approach is not as sensitive as an independent collection of swabs for RT-PCR testing using dedicated transport media. As such, the sensitivity of Ag-RDTs may be overestimated in this project, but this would apply to each swab type. However, not all real-world settings with vaccinated individuals still aim to identify the presence of molecular viral genetic material. The use of RT-PCR in this case was to provide a traditional comparison for laboratory testing purposes^24^, that may not be relevant in a post-vaccine pragmatic setting^25^. It should be noted that the use of RTB for RT-PCR testing avoids the additional nasal and throat collections for Ag-RDT and molecular testing. This work showed both the throat swab alone and the combined nasal/throat swab were well accepted by participants; however, it is unclear whether the same level of participation or acceptability would occur if 4 swabs were required for evaluations (2 each for Ag-RDT and RT-PCR testing). Finally, as Omicron was the only SARS-CoV-2 lineage circulating at the time, whole genome sequencing was not performed on the nucleic acids extracted from each swab; however, all positive samples lacked amplification of the S-gene target which is consistent with, but not confirmatory of detection of Omicron^26^. This may be relevant as different variants may be optimally detected with sampling of different anatomical sites, and our results are only generalizable to this presumed Omicron with potential tropism differences compared to previously circulating SARS-CoV-2 lineages.

Overall, this work provides early evidence that combined throat and bilateral nasal swabs maximize the ability of Ag-RDTs to identify the Omicron variant. These types of projects are instrumental in testing anecdotal reports to inform the community practice and increase confidence for those making swabbing recommendations to various stakeholders and users, including asymptomatic people performing self Ag-RDTs.

## Data Availability

All data produced in the present work are contained in the manuscript.

## Acknowledgements

The authors are indebted to the community participants for this evaluation, and to the many Test to Protect volunteers and coaches who have worked countless hours throughout the pandemic to ensure the safety of their community and community members.

## Ethics

This project was deemed a quality initiative and was therefore exempt from review by the Nova Scotia Health Research Ethics Board (submission number 1027644). Specimens tested were obtained from consenting participants, and all data related were provided anonymized, de-identified, and were used solely with the intent to evaluate the performance characteristics of the different swab types for rapid antigen testing programs used in Nova Scotia.

## Author contributions

All authors were involved in the design, data acquisition, and data interpretation. GP drafted the initial manuscript, with all authors contributing, and agree with the content of the final version.

---

**Supplemental Table 1.**
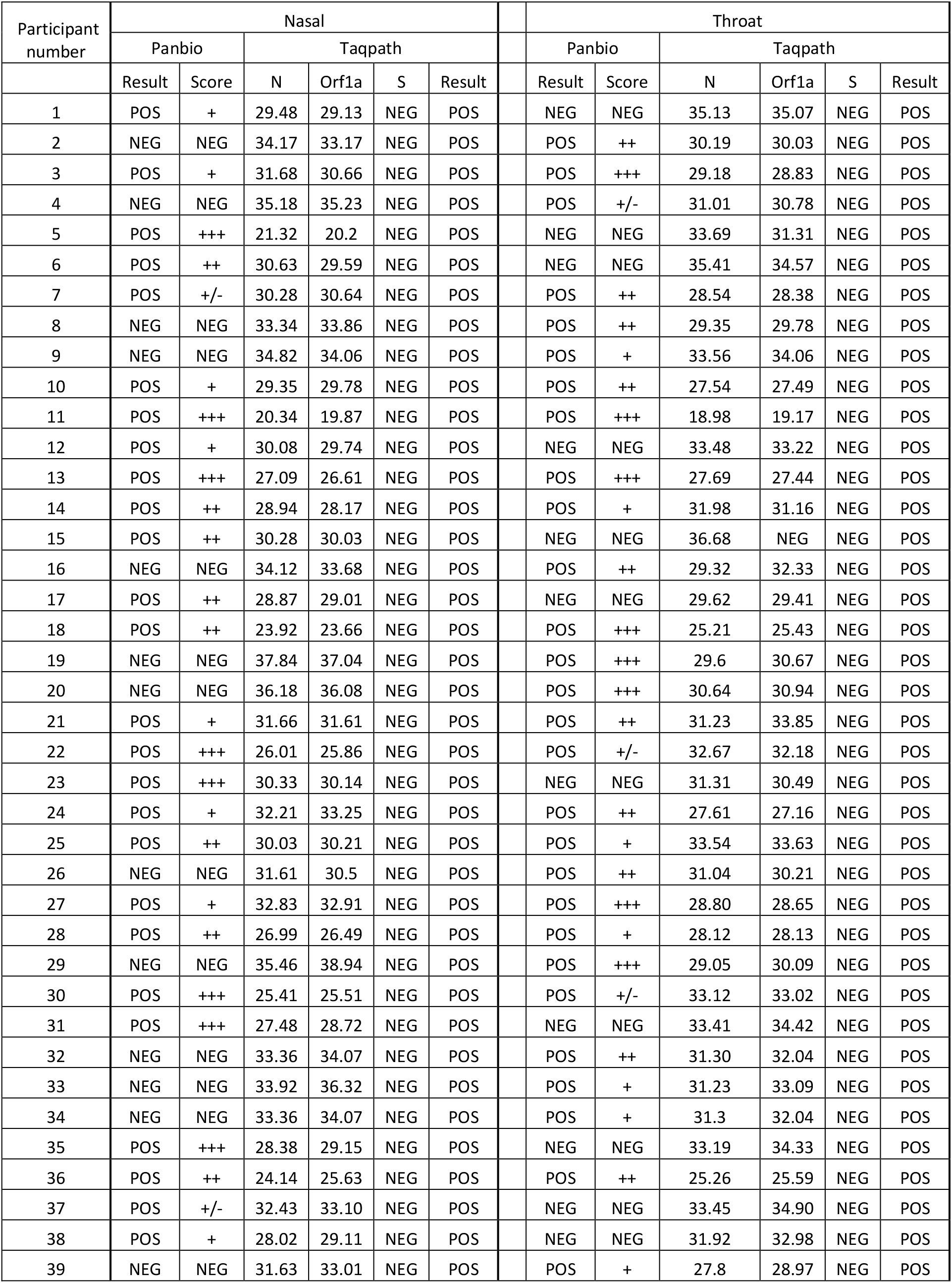
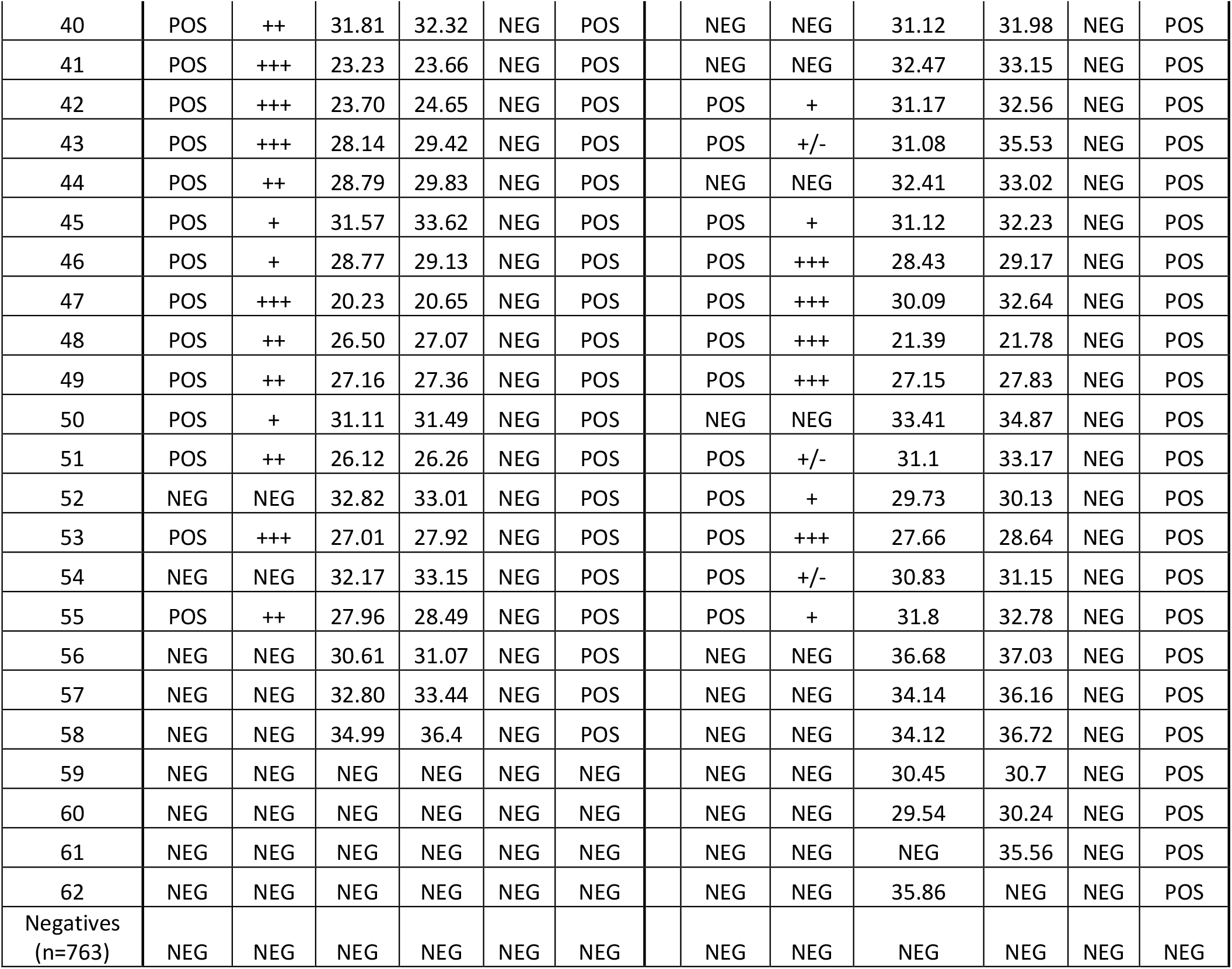
Nasal vs. Throat (Panbio)

| Participant number | Nasal |  |  |  |  |  | Throat |  |  |  |  |  |
| --- | --- | --- | --- | --- | --- | --- | --- | --- | --- | --- | --- | --- |
|  | Panbio |  | Taqpath |  |  |  | Panbio |  | Taqpath |  |  |  |
|  | Result | Score | N | Orf1a | S | Result | Result | Score | N | Orf1a | S | Result |
| 1 | POS | + | 29.48 | 29.13 | NEG | POS | NEG | NEG | 35.13 | 35.07 | NEG | POS |
| 2 | NEG | NEG | 34.17 | 33.17 | NEG | POS | POS | ++ | 30.19 | 30.03 | NEG | POS |
| 3 | POS | + | 31.68 | 30.66 | NEG | POS | POS | +++ | 29.18 | 28.83 | NEG | POS |
| 4 | NEG | NEG | 35.18 | 35.23 | NEG | POS | POS | +/- | 31.01 | 30.78 | NEG | POS |
| 5 | POS | +++ | 21.32 | 20.2 | NEG | POS | NEG | NEG | 33.69 | 31.31 | NEG | POS |
| 6 | POS | ++ | 30.63 | 29.59 | NEG | POS | NEG | NEG | 35.41 | 34.57 | NEG | POS |
| 7 | POS | +/- | 30.28 | 30.64 | NEG | POS | POS | ++ | 28.54 | 28.38 | NEG | POS |
| 8 | NEG | NEG | 33.34 | 33.86 | NEG | POS | POS | ++ | 29.35 | 29.78 | NEG | POS |
| 9 | NEG | NEG | 34.82 | 34.06 | NEG | POS | POS | + | 33.56 | 34.06 | NEG | POS |
| 10 | POS | + | 29.35 | 29.78 | NEG | POS | POS | ++ | 27.54 | 27.49 | NEG | POS |
| 11 | POS | +++ | 20.34 | 19.87 | NEG | POS | POS | +++ | 18.98 | 19.17 | NEG | POS |
| 12 | POS | + | 30.08 | 29.74 | NEG | POS | NEG | NEG | 33.48 | 33.22 | NEG | POS |
| 13 | POS | +++ | 27.09 | 26.61 | NEG | POS | POS | +++ | 27.69 | 27.44 | NEG | POS |
| 14 | POS | ++ | 28.94 | 28.17 | NEG | POS | POS | + | 31.98 | 31.16 | NEG | POS |
| 15 | POS | ++ | 30.28 | 30.03 | NEG | POS | NEG | NEG | 36.68 | NEG | NEG | POS |
| 16 | NEG | NEG | 34.12 | 33.68 | NEG | POS | POS | ++ | 29.32 | 32.33 | NEG | POS |
| 17 | POS | ++ | 28.87 | 29.01 | NEG | POS | NEG | NEG | 29.62 | 29.41 | NEG | POS |
| 18 | POS | ++ | 23.92 | 23.66 | NEG | POS | POS | +++ | 25.21 | 25.43 | NEG | POS |
| 19 | NEG | NEG | 37.84 | 37.04 | NEG | POS | POS | +++ | 29.6 | 30.67 | NEG | POS |
| 20 | NEG | NEG | 36.18 | 36.08 | NEG | POS | POS | +++ | 30.64 | 30.94 | NEG | POS |
| 21 | POS | + | 31.66 | 31.61 | NEG | POS | POS | ++ | 31.23 | 33.85 | NEG | POS |
| 22 | POS | +++ | 26.01 | 25.86 | NEG | POS | POS | +/- | 32.67 | 32.18 | NEG | POS |
| 23 | POS | +++ | 30.33 | 30.14 | NEG | POS | NEG | NEG | 31.31 | 30.49 | NEG | POS |
| 24 | POS | + | 32.21 | 33.25 | NEG | POS | POS | ++ | 27.61 | 27.16 | NEG | POS |
| 25 | POS | ++ | 30.03 | 30.21 | NEG | POS | POS | + | 33.54 | 33.63 | NEG | POS |
| 26 | NEG | NEG | 31.61 | 30.5 | NEG | POS | POS | ++ | 31.04 | 30.21 | NEG | POS |
| 27 | POS | + | 32.83 | 32.91 | NEG | POS | POS | +++ | 28.80 | 28.65 | NEG | POS |
| 28 | POS | ++ | 26.99 | 26.49 | NEG | POS | POS | + | 28.12 | 28.13 | NEG | POS |
| 29 | NEG | NEG | 35.46 | 38.94 | NEG | POS | POS | +++ | 29.05 | 30.09 | NEG | POS |
| 30 | POS | +++ | 25.41 | 25.51 | NEG | POS | POS | +/- | 33.12 | 33.02 | NEG | POS |
| 31 | POS | +++ | 27.48 | 28.72 | NEG | POS | NEG | NEG | 33.41 | 34.42 | NEG | POS |
| 32 | NEG | NEG | 33.36 | 34.07 | NEG | POS | POS | ++ | 31.30 | 32.04 | NEG | POS |
| 33 | NEG | NEG | 33.92 | 36.32 | NEG | POS | POS | + | 31.23 | 33.09 | NEG | POS |
| 34 | NEG | NEG | 33.36 | 34.07 | NEG | POS | POS | + | 31.3 | 32.04 | NEG | POS |
| 35 | POS | +++ | 28.38 | 29.15 | NEG | POS | NEG | NEG | 33.19 | 34.33 | NEG | POS |
| 36 | POS | ++ | 24.14 | 25.63 | NEG | POS | POS | ++ | 25.26 | 25.59 | NEG | POS |
| 37 | POS | +/- | 32.43 | 33.10 | NEG | POS | NEG | NEG | 33.45 | 34.90 | NEG | POS |
| 38 | POS | + | 28.02 | 29.11 | NEG | POS | NEG | NEG | 31.92 | 32.98 | NEG | POS |
| 39 | NEG | NEG | 31.63 | 33.01 | NEG | POS | POS | + | 27.8 | 28.97 | NEG | POS |

|  |  |  |  |  |  |  |  |  |  |  |  |  |  |
| --- | --- | --- | --- | --- | --- | --- | --- | --- | --- | --- | --- | --- | --- |
| 40 | POS | ++ | 31.81 | 32.32 | NEG | POS |  | NEG | NEG | 31.12 | 31.98 | NEG | POS |
| 41 | POS | +++ | 23.23 | 23.66 | NEG | POS |  | NEG | NEG | 32.47 | 33.15 | NEG | POS |
| 42 | POS | +++ | 23.70 | 24.65 | NEG | POS |  | POS | + | 31.17 | 32.56 | NEG | POS |
| 43 | POS | +++ | 28.14 | 29.42 | NEG | POS |  | POS | +/- | 31.08 | 35.53 | NEG | POS |
| 44 | POS | ++ | 28.79 | 29.83 | NEG | POS |  | NEG | NEG | 32.41 | 33.02 | NEG | POS |
| 45 | POS | + | 31.57 | 33.62 | NEG | POS |  | POS | + | 31.12 | 32.23 | NEG | POS |
| 46 | POS | + | 28.77 | 29.13 | NEG | POS |  | POS | +++ | 28.43 | 29.17 | NEG | POS |
| 47 | POS | +++ | 20.23 | 20.65 | NEG | POS |  | POS | +++ | 30.09 | 32.64 | NEG | POS |
| 48 | POS | ++ | 26.50 | 27.07 | NEG | POS |  | POS | +++ | 21.39 | 21.78 | NEG | POS |
| 49 | POS | ++ | 27.16 | 27.36 | NEG | POS |  | POS | +++ | 27.15 | 27.83 | NEG | POS |
| 50 | POS | + | 31.11 | 31.49 | NEG | POS |  | NEG | NEG | 33.41 | 34.87 | NEG | POS |
| 51 | POS | ++ | 26.12 | 26.26 | NEG | POS |  | POS | +/- | 31.1 | 33.17 | NEG | POS |
| 52 | NEG | NEG | 32.82 | 33.01 | NEG | POS |  | POS | + | 29.73 | 30.13 | NEG | POS |
| 53 | POS | +++ | 27.01 | 27.92 | NEG | POS |  | POS | +++ | 27.66 | 28.64 | NEG | POS |
| 54 | NEG | NEG | 32.17 | 33.15 | NEG | POS |  | POS | +/- | 30.83 | 31.15 | NEG | POS |
| 55 | POS | ++ | 27.96 | 28.49 | NEG | POS |  | POS | + | 31.8 | 32.78 | NEG | POS |
| 56 | NEG | NEG | 30.61 | 31.07 | NEG | POS |  | NEG | NEG | 36.68 | 37.03 | NEG | POS |
| 57 | NEG | NEG | 32.80 | 33.44 | NEG | POS |  | NEG | NEG | 34.14 | 36.16 | NEG | POS |
| 58 | NEG | NEG | 34.99 | 36.4 | NEG | POS |  | NEG | NEG | 34.12 | 36.72 | NEG | POS |
| 59 | NEG | NEG | NEG | NEG | NEG | NEG |  | NEG | NEG | 30.45 | 30.7 | NEG | POS |
| 60 | NEG | NEG | NEG | NEG | NEG | NEG |  | NEG | NEG | 29.54 | 30.24 | NEG | POS |
| 61 | NEG | NEG | NEG | NEG | NEG | NEG |  | NEG | NEG | NEG | 35.56 | NEG | POS |
| 62 | NEG | NEG | NEG | NEG | NEG | NEG |  | NEG | NEG | 35.86 | NEG | NEG | POS |
| Negatives<br>(n=763) | NEG | NEG | NEG | NEG | NEG | NEG |  | NEG | NEG | NEG | NEG | NEG | NEG |

**Supplemental Table 2.**
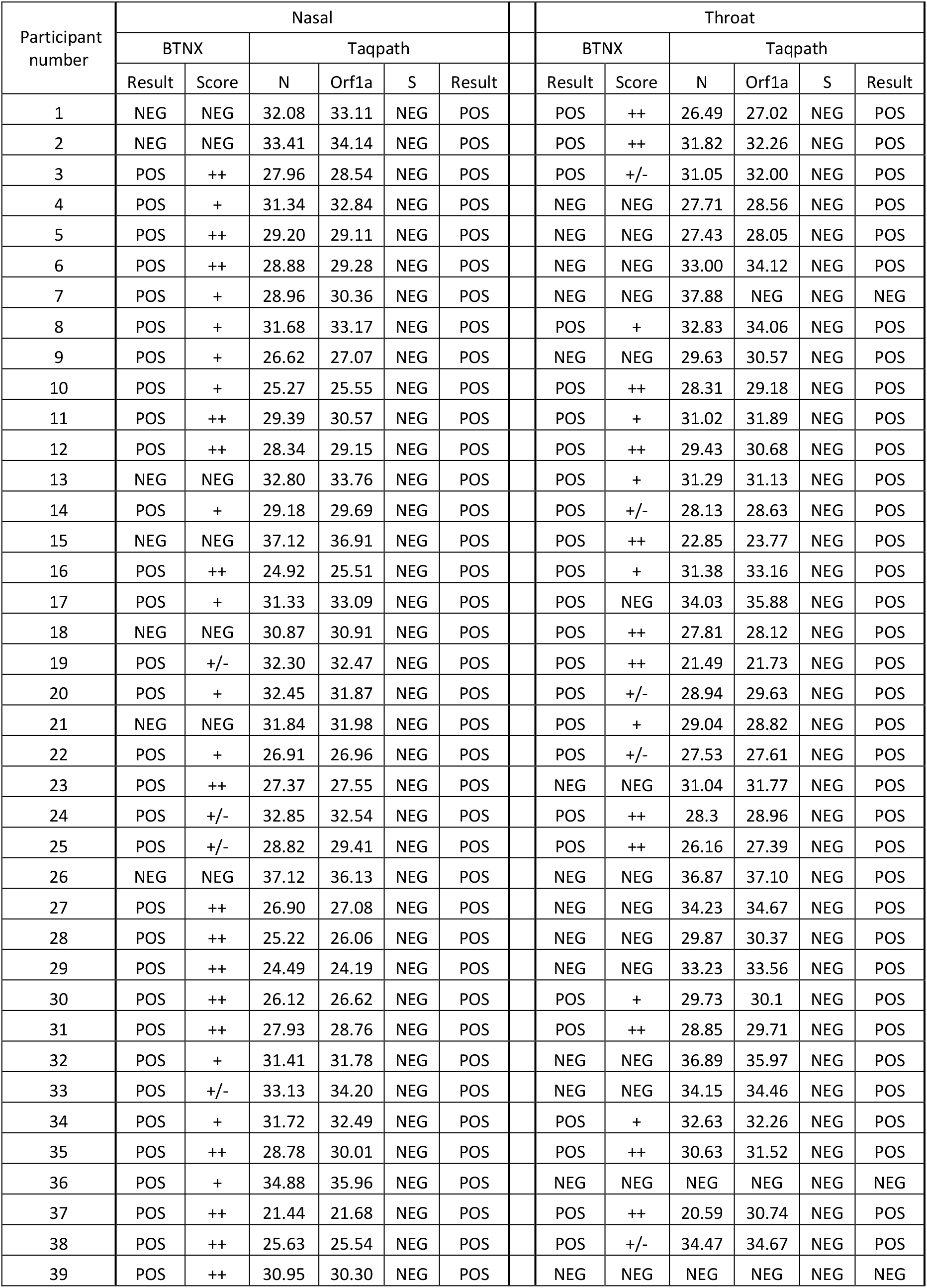
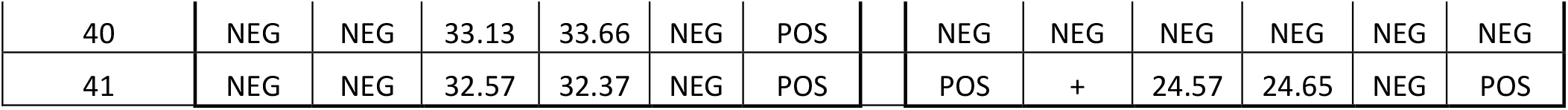
Nasal vs. Throat (BTNX)

| Participant number | Nasal |  |  |  |  |  | Throat |  |  |  |  |  |
| --- | --- | --- | --- | --- | --- | --- | --- | --- | --- | --- | --- | --- |
|  | BTNX |  | Taqpath |  |  |  | BTNX |  | Taqpath |  |  |  |
|  | Result | Score | N | Orf1a | S | Result | Result | Score | N | Orf1a | S | Result |
| 1 | NEG | NEG | 32.08 | 33.11 | NEG | POS | POS | ++ | 26.49 | 27.02 | NEG | POS |
| 2 | NEG | NEG | 33.41 | 34.14 | NEG | POS | POS | ++ | 31.82 | 32.26 | NEG | POS |
| 3 | POS | ++ | 27.96 | 28.54 | NEG | POS | POS | +/- | 31.05 | 32.00 | NEG | POS |
| 4 | POS | + | 31.34 | 32.84 | NEG | POS | NEG | NEG | 27.71 | 28.56 | NEG | POS |
| 5 | POS | ++ | 29.20 | 29.11 | NEG | POS | NEG | NEG | 27.43 | 28.05 | NEG | POS |
| 6 | POS | ++ | 28.88 | 29.28 | NEG | POS | NEG | NEG | 33.00 | 34.12 | NEG | POS |
| 7 | POS | + | 28.96 | 30.36 | NEG | POS | NEG | NEG | 37.88 | NEG | NEG | NEG |
| 8 | POS | + | 31.68 | 33.17 | NEG | POS | POS | + | 32.83 | 34.06 | NEG | POS |
| 9 | POS | + | 26.62 | 27.07 | NEG | POS | NEG | NEG | 29.63 | 30.57 | NEG | POS |
| 10 | POS | + | 25.27 | 25.55 | NEG | POS | POS | ++ | 28.31 | 29.18 | NEG | POS |
| 11 | POS | ++ | 29.39 | 30.57 | NEG | POS | POS | + | 31.02 | 31.89 | NEG | POS |
| 12 | POS | ++ | 28.34 | 29.15 | NEG | POS | POS | ++ | 29.43 | 30.68 | NEG | POS |
| 13 | NEG | NEG | 32.80 | 33.76 | NEG | POS | POS | + | 31.29 | 31.13 | NEG | POS |
| 14 | POS | + | 29.18 | 29.69 | NEG | POS | POS | +/- | 28.13 | 28.63 | NEG | POS |
| 15 | NEG | NEG | 37.12 | 36.91 | NEG | POS | POS | ++ | 22.85 | 23.77 | NEG | POS |
| 16 | POS | ++ | 24.92 | 25.51 | NEG | POS | POS | + | 31.38 | 33.16 | NEG | POS |
| 17 | POS | + | 31.33 | 33.09 | NEG | POS | POS | NEG | 34.03 | 35.88 | NEG | POS |
| 18 | NEG | NEG | 30.87 | 30.91 | NEG | POS | POS | ++ | 27.81 | 28.12 | NEG | POS |
| 19 | POS | +/- | 32.30 | 32.47 | NEG | POS | POS | ++ | 21.49 | 21.73 | NEG | POS |
| 20 | POS | + | 32.45 | 31.87 | NEG | POS | POS | +/- | 28.94 | 29.63 | NEG | POS |
| 21 | NEG | NEG | 31.84 | 31.98 | NEG | POS | POS | + | 29.04 | 28.82 | NEG | POS |
| 22 | POS | + | 26.91 | 26.96 | NEG | POS | POS | +/- | 27.53 | 27.61 | NEG | POS |
| 23 | POS | ++ | 27.37 | 27.55 | NEG | POS | NEG | NEG | 31.04 | 31.77 | NEG | POS |
| 24 | POS | +/- | 32.85 | 32.54 | NEG | POS | POS | ++ | 28.3 | 28.96 | NEG | POS |
| 25 | POS | +/- | 28.82 | 29.41 | NEG | POS | POS | ++ | 26.16 | 27.39 | NEG | POS |
| 26 | NEG | NEG | 37.12 | 36.13 | NEG | POS | NEG | NEG | 36.87 | 37.10 | NEG | POS |
| 27 | POS | ++ | 26.90 | 27.08 | NEG | POS | NEG | NEG | 34.23 | 34.67 | NEG | POS |
| 28 | POS | ++ | 25.22 | 26.06 | NEG | POS | NEG | NEG | 29.87 | 30.37 | NEG | POS |
| 29 | POS | ++ | 24.49 | 24.19 | NEG | POS | NEG | NEG | 33.23 | 33.56 | NEG | POS |
| 30 | POS | ++ | 26.12 | 26.62 | NEG | POS | POS | + | 29.73 | 30.1 | NEG | POS |
| 31 | POS | ++ | 27.93 | 28.76 | NEG | POS | POS | ++ | 28.85 | 29.71 | NEG | POS |
| 32 | POS | + | 31.41 | 31.78 | NEG | POS | NEG | NEG | 36.89 | 35.97 | NEG | POS |
| 33 | POS | +/- | 33.13 | 34.20 | NEG | POS | NEG | NEG | 34.15 | 34.46 | NEG | POS |
| 34 | POS | + | 31.72 | 32.49 | NEG | POS | POS | + | 32.63 | 32.26 | NEG | POS |
| 35 | POS | ++ | 28.78 | 30.01 | NEG | POS | POS | ++ | 30.63 | 31.52 | NEG | POS |
| 36 | POS | + | 34.88 | 35.96 | NEG | POS | NEG | NEG | NEG | NEG | NEG | NEG |
| 37 | POS | ++ | 21.44 | 21.68 | NEG | POS | POS | ++ | 20.59 | 30.74 | NEG | POS |
| 38 | POS | ++ | 25.63 | 25.54 | NEG | POS | POS | +/- | 34.47 | 34.67 | NEG | POS |
| 39 | POS | ++ | 30.95 | 30.30 | NEG | POS | NEG | NEG | NEG | NEG | NEG | NEG |

|  |  |  |  |  |  |  |  |  |  |  |  |  |  |
| --- | --- | --- | --- | --- | --- | --- | --- | --- | --- | --- | --- | --- | --- |
| 40 | NEG | NEG | 33.13 | 33.66 | NEG | POS |  | NEG | NEG | NEG | NEG | NEG | NEG |
| 41 | NEG | NEG | 32.57 | 32.37 | NEG | POS |  | POS | + | 24.57 | 24.65 | NEG | POS |

**Supplemental Table 3.**
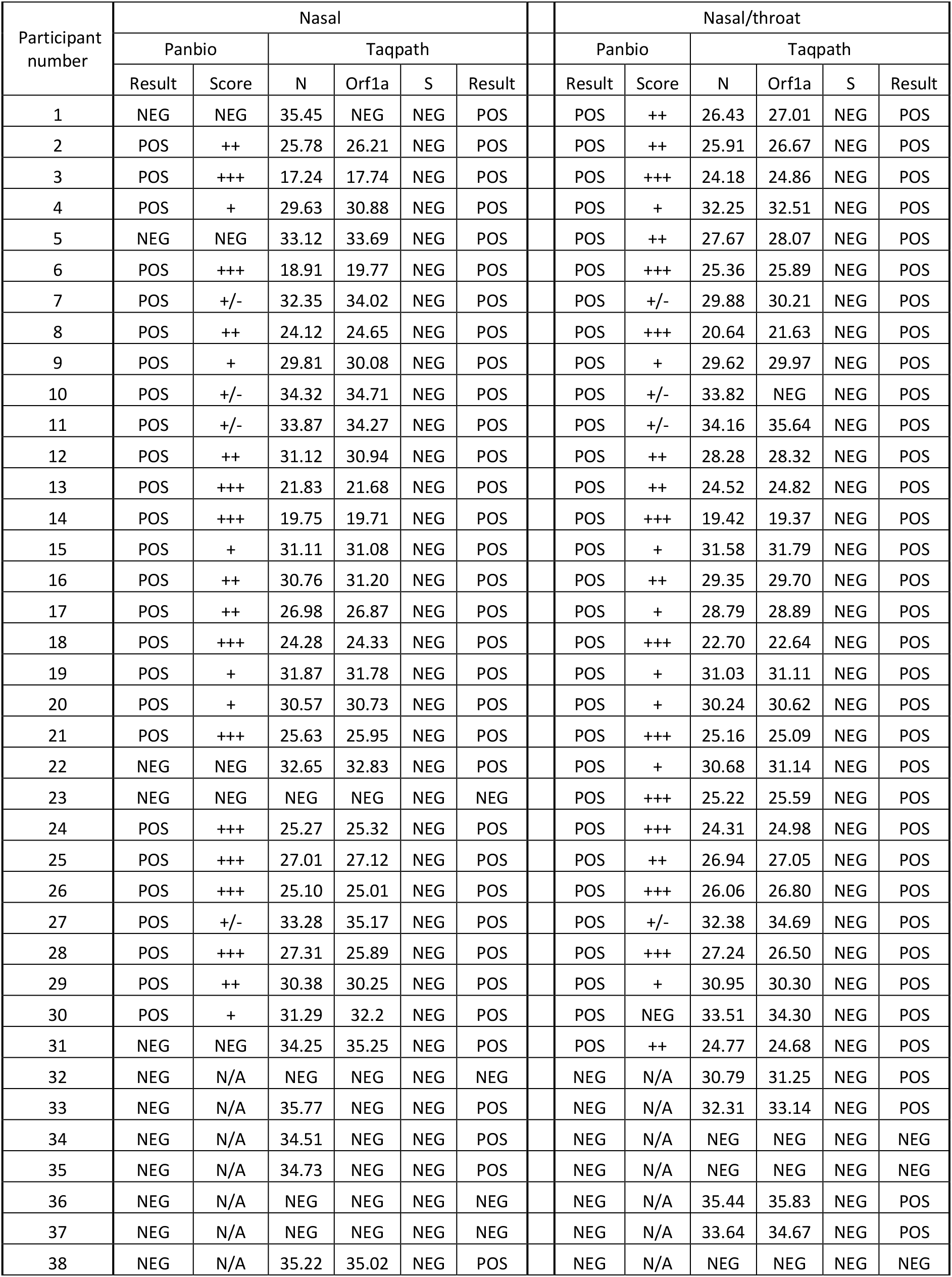
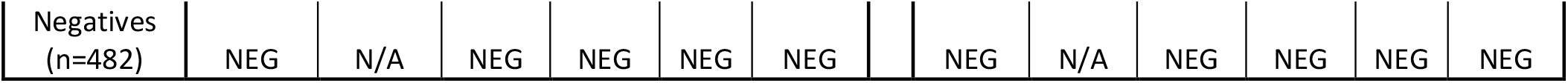
Nasal sample vs. Throat + Nasal combo swab (Panbio)

| Participant number | Nasal |  |  |  |  |  | Nasal/throat |  |  |  |  |  |
| --- | --- | --- | --- | --- | --- | --- | --- | --- | --- | --- | --- | --- |
|  | Panbio |  | Taqpath |  |  |  | Panbio |  | Taqpath |  |  |  |
|  | Result | Score | N | Orf1a | S | Result | Result | Score | N | Orf1a | S | Result |
| 1 | NEG | NEG | 35.45 | NEG | NEG | POS | POS | ++ | 26.43 | 27.01 | NEG | POS |
| 2 | POS | ++ | 25.78 | 26.21 | NEG | POS | POS | ++ | 25.91 | 26.67 | NEG | POS |
| 3 | POS | +++ | 17.24 | 17.74 | NEG | POS | POS | +++ | 24.18 | 24.86 | NEG | POS |
| 4 | POS | + | 29.63 | 30.88 | NEG | POS | POS | + | 32.25 | 32.51 | NEG | POS |
| 5 | NEG | NEG | 33.12 | 33.69 | NEG | POS | POS | ++ | 27.67 | 28.07 | NEG | POS |
| 6 | POS | +++ | 18.91 | 19.77 | NEG | POS | POS | +++ | 25.36 | 25.89 | NEG | POS |
| 7 | POS | +/- | 32.35 | 34.02 | NEG | POS | POS | +/- | 29.88 | 30.21 | NEG | POS |
| 8 | POS | ++ | 24.12 | 24.65 | NEG | POS | POS | +++ | 20.64 | 21.63 | NEG | POS |
| 9 | POS | + | 29.81 | 30.08 | NEG | POS | POS | + | 29.62 | 29.97 | NEG | POS |
| 10 | POS | +/- | 34.32 | 34.71 | NEG | POS | POS | +/- | 33.82 | NEG | NEG | POS |
| 11 | POS | +/- | 33.87 | 34.27 | NEG | POS | POS | +/- | 34.16 | 35.64 | NEG | POS |
| 12 | POS | ++ | 31.12 | 30.94 | NEG | POS | POS | ++ | 28.28 | 28.32 | NEG | POS |
| 13 | POS | +++ | 21.83 | 21.68 | NEG | POS | POS | ++ | 24.52 | 24.82 | NEG | POS |
| 14 | POS | +++ | 19.75 | 19.71 | NEG | POS | POS | +++ | 19.42 | 19.37 | NEG | POS |
| 15 | POS | + | 31.11 | 31.08 | NEG | POS | POS | + | 31.58 | 31.79 | NEG | POS |
| 16 | POS | ++ | 30.76 | 31.20 | NEG | POS | POS | ++ | 29.35 | 29.70 | NEG | POS |
| 17 | POS | ++ | 26.98 | 26.87 | NEG | POS | POS | + | 28.79 | 28.89 | NEG | POS |
| 18 | POS | +++ | 24.28 | 24.33 | NEG | POS | POS | +++ | 22.70 | 22.64 | NEG | POS |
| 19 | POS | + | 31.87 | 31.78 | NEG | POS | POS | + | 31.03 | 31.11 | NEG | POS |
| 20 | POS | + | 30.57 | 30.73 | NEG | POS | POS | + | 30.24 | 30.62 | NEG | POS |
| 21 | POS | +++ | 25.63 | 25.95 | NEG | POS | POS | +++ | 25.16 | 25.09 | NEG | POS |
| 22 | NEG | NEG | 32.65 | 32.83 | NEG | POS | POS | + | 30.68 | 31.14 | NEG | POS |
| 23 | NEG | NEG | NEG | NEG | NEG | NEG | POS | +++ | 25.22 | 25.59 | NEG | POS |
| 24 | POS | +++ | 25.27 | 25.32 | NEG | POS | POS | +++ | 24.31 | 24.98 | NEG | POS |
| 25 | POS | +++ | 27.01 | 27.12 | NEG | POS | POS | ++ | 26.94 | 27.05 | NEG | POS |
| 26 | POS | +++ | 25.10 | 25.01 | NEG | POS | POS | +++ | 26.06 | 26.80 | NEG | POS |
| 27 | POS | +/- | 33.28 | 35.17 | NEG | POS | POS | +/- | 32.38 | 34.69 | NEG | POS |
| 28 | POS | +++ | 27.31 | 25.89 | NEG | POS | POS | +++ | 27.24 | 26.50 | NEG | POS |
| 29 | POS | ++ | 30.38 | 30.25 | NEG | POS | POS | + | 30.95 | 30.30 | NEG | POS |
| 30 | POS | + | 31.29 | 32.2 | NEG | POS | POS | NEG | 33.51 | 34.30 | NEG | POS |
| 31 | NEG | NEG | 34.25 | 35.25 | NEG | POS | POS | ++ | 24.77 | 24.68 | NEG | POS |
| 32 | NEG | N/A | NEG | NEG | NEG | NEG | NEG | N/A | 30.79 | 31.25 | NEG | POS |
| 33 | NEG | N/A | 35.77 | NEG | NEG | POS | NEG | N/A | 32.31 | 33.14 | NEG | POS |
| 34 | NEG | N/A | 34.51 | NEG | NEG | POS | NEG | N/A | NEG | NEG | NEG | NEG |
| 35 | NEG | N/A | 34.73 | NEG | NEG | POS | NEG | N/A | NEG | NEG | NEG | NEG |
| 36 | NEG | N/A | NEG | NEG | NEG | NEG | NEG | N/A | 35.44 | 35.83 | NEG | POS |
| 37 | NEG | N/A | NEG | NEG | NEG | NEG | NEG | N/A | 33.64 | 34.67 | NEG | POS |
| 38 | NEG | N/A | 35.22 | 35.02 | NEG | POS | NEG | N/A | NEG | NEG | NEG | NEG |

|  |  |  |  |  |  |  |  |  |  |  |  |  |  |
| --- | --- | --- | --- | --- | --- | --- | --- | --- | --- | --- | --- | --- | --- |
| Negatives<br>(n=482) | NEG | N/A | NEG | NEG | NEG | NEG |  | NEG | N/A | NEG | NEG | NEG | NEG |

